# Sex-specific risk loci and modified *MEF2C* expression in ALS

**DOI:** 10.1101/2024.05.25.24307829

**Authors:** Ross P Byrne, Wouter van Rheenen, Tomás DS Gomes, Ciarán M Kelly, Emrah Kaçar, Project MinE ALS GWAS Consortium, International ALS/FTD Genomics Consortium, Ahmad Al Khleifat, Alfredo Iacoangeli, Ammar Al-Chalabi, Jan H Veldink, Russell L McLaughlin

**Affiliations:** Complex Trait Genomics Laboratory, Smurfit Institute of Genetics, Trinity College Dublin, D02 PN40, Ireland; Department of Neurology, Brain Center Rudolf Magnus, University Medical Center Utrecht, Utrecht, the Netherlands; School of Pharmacy and Biomolecular Sciences, RCSI University of Medicine and Health Sciences, Dublin, Ireland; King’s College London, Maurice Wohl Clinical Neuroscience Institute, Department of Basic and Clinical Neuroscience, London SE5 9RX, United Kingdom; Perron Institute for Neurological and Translational Science, University of Western Australia Medical School, Perth, WA 6009, Australia; Department of Biostatistics and Health Informatics, King’s College London, London, United Kingdom; NIHR BRC SLAM NHS Foundation Trust, London, United Kingdom; King’s College Hospital, Denmark Hill, SE5 9RS London, UK

**Author notes:** To whom correspondence should be addressed: Ross P Byrne PhD, Complex Trait Genomics Laboratory, Smurfit Institute of Genetics, Trinity College Dublin, D02 PN40, Republic of Ireland. A full list of authors and their affiliations appears in Supplementary note 1. A full list of authors and their affiliations appears in Supplementary note 2.

## Abstract

Significantly more men develop amyotrophic lateral sclerosis (ALS) than women, and heritability is not uniform between male and female transmissions, together suggesting a role for sex in the genetic aetiology of the disease. We therefore performed sex-stratified genome-wide and transcriptome-wide analyses of ALS risk, identifying six novel sex-specific risk loci including *MEF2C*, which shows increased expression in female ALS motor neurones. X-chromosome analysis revealed an additional risk locus at *IL1RAPL2*.

---

We calculated that previously-reported^1^ differences in like-sexed and unlike-sexed parent-offspring transmission concordance for ALS implies a female-male genetic correlation of 62.8% (95% c.i. 48.0-73.2%), suggesting that some inherited ALS risk factors differ between women and men. To investigate this, we re-analysed the largest genome-wide association study (GWAS) of ALS^2^, splitting autosomal data by sex (*n*_female_ = 67,957; *n*_male_ = 65,692; Supplementary table 1), which revealed novel female-specific risk loci at *FGFRL1* and *MEF2C* and male-specific risk loci at *CCDC85A, RESP18, WIPI2* and *LUZP2* (Figure 1, Supplementary table 2). A further two previously-implicated loci (HLA locus and *SCFD1*) were only significantly associated with ALS risk in males, but did not meet our definition of sex-specific (p < 2.3 × 10^−7^ in one sex, corresponding to a false discovery rate < 1%, and p > 0.05 in the other sex). GWAS statistics for both sexes were modestly inflated (*λ*_GC_ = 1.03 in both; Supplementary figure 1) suggesting some polygenic risk, which was further supported by univariate LD score regression^3^ heritability estimates (*h*^2^_SNP_ (s.e.) = 2.8% (0.65) for females and 2.1% (0.42) for males), with no apparent role for confounding in GWAS inflation (LDSC intercepts < 1). Despite the identification of sex-specific loci, bivariate LD score regression^4^ estimated the genetic correlation between male and female autosomal GWAS statistics to be 99% (s.e. 16%; p=4.1×10^−10^), suggesting an overall shared polygenic component of ALS risk architecture between the two sexes. X chromosome data were available for a subset of individuals (*n*_female_ = 19,576; *n*_male_ = 21,991); these were analysed per sex then resulting statistics were meta-analysed, revealing one further novel ALS-associated risk locus at *IL1RAPL2* (Figure 1 Supplementary table 3).

**Figure 1.**
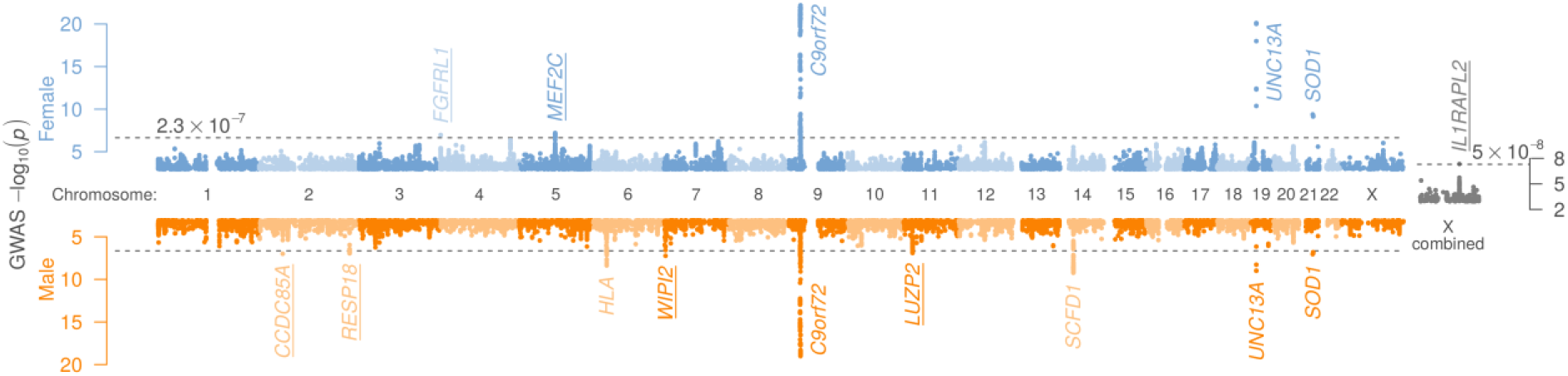
Sex-stratified genome-wide association study (GWAS) statistics for amyotrophic lateral sclerosis (ALS) risk, including sex-combined X-chromosome association statistics. For sex-stratified analyses, the p-value threshold to consider a locus as significantly associated with ALS corresponds to a false discovery rate of 1%. Combined X chromosome analysis uses the conventional genome-wide significance threshold of 5 × 10^−8^. Significant loci are annotated with their closest gene and for brevity, only loci where p < 0.001 are plotted. For the six novel sex-specific loci (underlined), cohort- and locus-level detail are provided in Supplementary figure 2-7.

To identify gene expression modified by ALS risk alleles, we performed a transcriptome-wide association study (TWAS) of sex-stratified GWAS results with expression data for 13 brain tissues in GTEx version 8. A previously-noted^5^ association between ALS risk and expression of *C9orf72* was observed in both sexes and modified expression of *MEF2C* and *RESP18* were associated with ALS risk in a strongly sex-specific manner (Figure 2A, Supplementary table 4). Of the other GWAS risk loci, only *SCFD1* was significantly associated by TWAS analysis in males, but this was not sex-specific (*p*_female_ = 1.81 × 10^−5^). Notably, independent analysis of motor neurones (MNs) derived from induced pluripotent stem cells (IPSCs) from ALS patients and controls^6^ revealed *MEF2C* expression to be significantly higher in female ALS patients than in female controls (*p*_female_ = 3×10^−4^, Figure 2B), but not significantly different between male ALS patients and male controls (*p*_male_ = 0.07).

**Figure 2.**
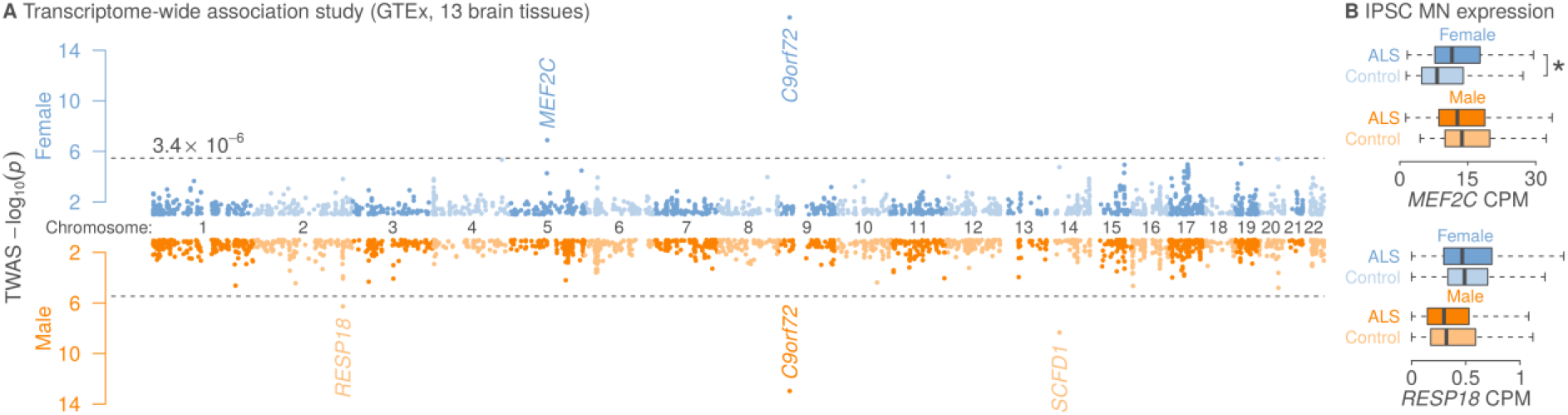
Sex-specific analysis of gene expression in amyotrophic lateral sclerosis (ALS). (**A**) Transcriptome-wide association study (TWAS) statistics using GWAS results from Figure 1 analysed with expression data from 13 brain tissues in GTEx version 8. Significance threshold is Bonferroni-corrected for 14,706 tests. Genes whose expression are significantly modified by ALS GWAS risk alleles are annotated and for brevity, only loci where p < 0.1 are plotted. (**B**) For TWAS loci identified as sex-specific (p < 3.4 × 10^−6^ in one sex; p > 0.05 in the other), boxplots show sex-stratified expression (counts per million, CPM) in an independent set of motor neurones (MNs) derived from induced pluripotent stem cells (IPSCs) generated from 471 ALS patients and 283 age-matched controls in Answer ALS^**6**^. * p < 8.3 × 10^−3^ (logistic regression, corrected for 6 tests).

*MEF2C* is a transcription factor involved in development of the mammalian neocortex, and has been associated with Alzheimer’s and Parkinson’s diseases, along with several neuropsychiatric traits. As ALS risk modulated by *MEF2C* expression likely involves its transcriptional targets, we reasoned that genetic variation present in target genes may also confer risk for ALS. We therefore performed stratified LD score regression^7^ to identify partitioned heritability enrichment for ALS in a set of 1,055 genes either up- or downregulated in *Mef2c* conditional knockout mice^8,9^, revealing 2.4-fold heritability enrichment in human orthologues of downregulated genes in female ALS GWAS statistics (*p* = 2.6 × 10^−3^) and a statistically insignificant heritability depletion in upregulated targets (*p* = 0.56). No significant heritability enrichment was observed in male ALS GWAS statistics (Supplementary table 5).

It is possible that *MEF2C* transcriptomic results partially explain the sex ratio in ALS^10^: although dysregulation of *MEF2C* targets has previously been implicated in ALS in sex-agnostic analyses^11^, our data suggest that lower expression in female motor neurones is protective against ALS, and increased expression to the range observed in men (who are generally at higher risk) increases risk in women. It is also worth noting that the IPSC MN system is isolated from endocrine differences between men and women, so sex differences are likely due to cell-autonomous regulation or sex-divergent epigenetic cellular identity established over the lifetime of the individual. This contrasts with *RESP18*, whose expression in MNs was not significantly different between cases and controls of either sex, despite its association by TWAS in men. As *RESP18* encodes a secreted neuroendocrine protein, it is not necessarily expected that IPSC MN expression in isolation would differ between ALS cases and controls. However, MN expression was lower in men overall than in women (p = 1.09 × 10^−9^), and female rats have previously been shown to express twice the level of RESP18 protein in plasma compared to male rats^12^, consistent with a possible role for sex in determining its contribution to neurodegeneration. In support of this, *Resp18* knockout impairs locomotor activity and motor coordination but appears to protect against behavioural impairment in a chemically-induced model of Parkinson’s disease in male mice^13^.

Although gene expression changes at remaining novel autosomal GWAS loci (*CCDC85A, FGFRL1, WIPI2* and *LUZP2*) were not significantly associated with ALS by TWAS, these genes also have biological roles that may contribute to candidate disease mechanisms in ALS when their risk alleles are inherited. *CCDC85A* is highly expressed in the frontal cortex, associating with synaptic function and interactions between neurones and astrocytes. The *FGFRL1* locus also includes *IDUA*, which has been associated with Alzheimer’s disease^14^, and *GAK*, which was previously associated with Parkinson’s disease^15^ and implicated as a shared risk locus through colocalization analysis in sex-agnostic ALS GWAS^2^. *WIPI2* is involved in autophagy (a key process in ALS pathophysiology), and interacts with ALS-related proteins TBK1 and optineurin. *LUZP2* has recently been associated with age at onset of Alzheimer’s disease^16^. Finally, *IL1RAPL2* was revealed by X chromosome analysis, which has previously been overlooked in ALS genomics. This encodes a central nervous system-specific protein which is part of the interactome of interleukin-2, a cytokine currently under clinical trial investigating its potential neuroprotective effect in ALS^17^. Stratification of trial results by rs17332232 genotype may reveal a pharmacogenomic element to any observed therapeutic benefit.

## Methods

### Pedigree-based genetic correlation estimation

Heritability estimates from like-sexed (mother-daughter and father-son) and unlike-sexed (mother-son and father-daughter) parent-offspring pairings were obtained from Ryan et al (2019)^1^ to estimate genetic correlation in ALS liability between sexes. Falconer (1967)^18^ expresses this as

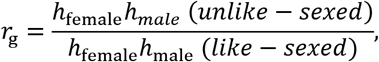

where *r*_g_ is the genetic correlation between sexes and *h*_female_ and *h*_male_ are the square roots of heritability in female and male cohorts, respectively. Confidence intervals were calculated using the upper and lower bounds of the 95% confidence intervals from the source study in the numerator and denominator, respectively for the upper bound, and *vice versa* for the lower bound.

### Sex-stratified genome-wide association study

Genotype data were obtained from the European component the ALS GWAS presented by van Rheenen et al. (2021)^2^. Only variants and individuals passing quality control (QC) described in the original publication were retained for analysis. The source study subdivides data into six strata representing similar genotyping platforms; these same strata were used and further subdivided into male-only and female-only subsets based on reported sex (Supplementary table 1). One sample from any pair with a coefficient of relatedness greater than 0.125 (calculated over 150k SNPs) between the male and female subsets of each stratum was removed to avoid potential bias in analyses comparing the two sex-specific GWAS subsets (e.g. genetic correlation analysis). Samples with relatedness within each stratum were retained; these are appropriately handled by SAIGE v0.39^19^, which we used to calculate association statistics per stratum. Firstly, we fit a null logistic mixed model in a leave-one chromosome out scheme per stratum using the first 20 principal components as covariates. For this model we used a subset of high-quality (INFO > 0.95) SNPs which were LD pruned using PLINK 1.9 (--indep-pairwise 50 25 0.05 and high LD regions removed: https://genome.sph.umich.edu/wiki/Regions_of_high_linkage_disequilibrium_(LD)). Next we ran a SNP-wise logistic mixed model implementing a saddlepoint approximation on all hard called genotypes in a leave one chromosome out scheme. Resulting sex-specific association statistics were combined across strata using inverse variance weighted meta-analysis implemented in METAL^20^. Associated loci were classified as sex-specific if significantly associated with ALS in one sex after stringent correction for multiple testing (1% false discovery rate; Benjamini-Hochberg) but not even nominally associated in the other sex (*p* > 0.05).

To estimate SNP-based heritability we ran univariate LD score regression on sex-specific GWAS summary statistics, using pre-calculated LD scores from the European subsample of the 1000 Genomes Project and considering only SNPs included in HapMap phase 3. Heritability estimates were converted to the liability scale using sex-specific lifetime risk estimates for ALS^1^ (1 in 436 for women; 1 in 347 for men) as prevalence. The sum of effective sample sizes (i.e. the effective sample size of an equivalent GWAS with a 50:50 case control split) was used in place of total N, and sample prevalence was set at 0.5 following evidence that using total N and sample prevalence downwardly biases heritability estimates when GWAS results from meta-analysis of several cohorts with different ascertainment^21^. Effective sample size was calculated as 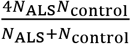 in each stratum and summed across strata.

### X chromosome analysis

Previously unanalysed X chromosome data for 41,567 samples (15,146 cases and 26,421 controls totalling 19,576 women and 21,991 men), compiled from two recent ALS GWAS cohorts^2,22^, were analysed to identify genetic associations between ALS and X-chromosome variants. Only samples passing QC in the base papers with associated X chromosome data were retained. This dataset was analysed separately from the autosomal GWAS datasets due to both the different sample composition (X chromosome data were unavailable for many samples) and different QC and analysis strategies needed to perform robust X chromosome association analysis.

Data from the raw cohorts were mapped to GRCh37 using liftOver and for each cohort we performed an initial pre-imputation QC. Briefly, we split cohorts by sex, then for each we removed: i) SNPs with missingness > 0.02 in either sex; ii) SNPs with minor allele frequencies (MAF) < 0.01 in either sex; iii) SNPs showing significant deviation from Hardy-Weinberg equilibrium (HWE; *p* < 1 × 10^−4^); iv) strand ambiguous AT or GC SNPs; v) SNPs showing heterozygous calls on the X-chromosome in men. Cohorts were then merged into 28 genotyping platform- and country-matched strata which were balanced for case-control ratio following the scheme of the source papers^2,22^ and subject to a second round of QC, removing SNPs per stratum with: i) significant deviation from HWE (*p* < 1 × 10^−5^); ii) missingness > 0.1; iii) MAF < 0.01. Finally, one sample from any pair with a coefficient of relatedness greater than 0.125 was removed.

For imputation, input VCF files aligned to the Haplotype Reference Consortium (HRC) reference allele were prepared following the Michigan Imputation Server best practices guidelines. Imputation was then carried out per stratum on the Michigan imputation server using the HRC (version r1.1 2016) as a reference panel, Eagle 2.4 for phasing and Minmac 4 for imputation. Following imputation, SNPs with an Rsq value of 0.3 or greater were retained and subjected to a final round of QC, including removal of SNPs: i) significantly deviating from HWE (*p* < 1 × 10^−5^) per stratum; ii) with missingness > 0.05 in the whole dataset; iii) with MAF < 0.01 in the whole dataset.

For association testing, strata were merged using plink, male genotypes were recoded as 0/1 and the software XWAS^23^ was run using a sex-stratified model with 20 principal components calculated from autosomal SNPs as covariates. Test statistics for males and females were combined across sexes using Fisher’s method (`--fisher`), which does not make assumptions about X inactivation, simplifying interpretation.

### Transcriptomic analyses

We carried out a transcriptome-wide association study (TWAS) using FUSION^24^ on the summary statistics from the male and female GWAS for ALS to test for association between gene expression and ALS in 13 brain tissues from GTEx version 8. For this analysis we used the European subset of the 1000 Genomes Project as an LD reference panel and precomputed weights from all GTEx samples (http://gusevlab.org/projects/fusion/). Resulting p-values per gene were aggregated across brain tissues using the SCAT method developed by Xiao et al. (2020)^5^.

To analyse sex-specific TWAS hits in IPSC-derived motor neurones, raw transcriptomic count data for 773 samples were downloaded from the Answer ALS data portal (version 6.0; January 2024). Definite ALS cases and controls were extracted based on clinical metadata, excluding non-ALS MND and asymptomatic carriers. Count data were normalised and converted to counts per million (CPM) using the edgeR package. Association between ALS and gene expression for *MEF2C* and *RESP18* was estimated using a logistic regression model in R and computed in the full dataset (471 ALS, 283 control) as well as male-stratified (294 ALS, 87 control) and female-stratified (177 ALS, 196 control) subsets of the data. A Bonferonni adjusted p-value threshold of P<0.0083 was used to correct for multiple testing (6 tests).

### Heritability enrichment of *MEF2C* targets

Stratified LD score regression^7^ was used to analyse heritability enrichment in a set of genes^9^ differentially expressed after *Mef2c* conditional knockout in developing mice^8^. This analysis was run using sex-stratified GWAS summary statistics and combined data from the European-only subset of the 2021 ALS GWAS^2^. An annotation was first made for SNPs in a 100 kb region surrounding each differentially-expressed gene. LD scores were calculated for this annotation using the European subset of the 1000 Genomes Project dataset and considering only HapMap 3 SNPs. The annotation was run against the baseline LD model.

## Supporting information

Supplementary_figs_tabs_text

Table with local IRBs

## Acknowledgements

This study was funded by the MND Association of England, Wales and Northern Ireland under grant number 879-791. Data used in the preparation of this article were obtained from the ANSWER ALS Data Portal (AALS-01184). For up-to-date information on the study, visit https://dataportal.answerals.org. This publication has emanated from research supported in part by a research grant from Science Foundation Ireland (SFI) under Grant Number 21/RC/10294 and co-funded under the European Regional Development Fund and by FutureNeuro industry partners. R.P.B. currently receives support from the MND Association (979-799). A.I. is funded by South London and Maudsley NHS Foundation Trust, MND Scotland, Motor Neurone Disease Association, National Institute for Health and Care Research, Spastic Paraplegia Foundation, Rosetrees Trust, Darby Rimmer MND Foundation, the Medical Research Council (UKRI), Alzheimer’s Research UK and LifeArc. AAK is funded by ALS Association Milton Safenowitz Research Fellowship (grant number22-PDF-609.DOI:10.52546/pc.gr.150909.), the Motor Neurone Disease Association (MNDA) Fellowship (Al Khleifat/Oct21/975-799), The Darby Rimmer Foundation, and The NIHR Maudsley Biomedical Research Centre. AAC is an NIHR Senior Investigator (NIHR202421). This is an EU Joint Programme - Neurodegenerative Disease Research (JPND) project. The project is supported through the following funding organisations under the aegis of JPND - www.jpnd.eu *(United Kingdom, Medical Research Council* (MR/L501529/1; MR/R024804/1) *and Economic and Social Research Council* (ES/L008238/1)) and through the Motor Neurone Disease Association, My Name’5 Doddie Foundation, and Alan Davidson Foundation. This study represents independent research part funded by the National Institute for Health Research (NIHR) Biomedical Research Centre at South London and Maudsley NHS Foundation Trust and King’s College London.

## Author contributions

R.L.McL. and R.P.B. conceived the study and drafted the manuscript. R.P.B. conducted the analyses with the assistance of T.D.S, C.M.K. and E.K. W.v.R., A.A-K., A.I., A.A-C. and J.H.V. supplied data for GWAS and XWAS analyses. All authors provided critical review of the manuscript.

## Data availability

The GWAS summary statistics generated in this study will be made publicly available upon publication in the NHGRI-EBI GWAS Catalog at https://www.ebi.ac.uk/gwas/ (for female, male and X chromosome meta-analyses, respectively) and through the Project MinE website (https://www.projectmine.com/research/download-data/). Individual level genotype data used in this study was originally published in van Rheenen et al 2021 which includes the following publicly available datasets: the Wellcome Trust Case Control Consortium (https://www.wtccc.org.uk/) and dbGaP datasets (phs000101.v3.p1, NIH Genome-Wide Association Studies of Amyotrophic Lateral Sclerosis; phs000126.v1.p1, CIDR: Genome Wide Association Study in Familial Parkinson Disease (PD); phs000196.v1.p1, Genome-Wide Association Study of Parkinson Disease: Genes and Environment; phs000344.v1.p1, Genome-Wide Association Study of Amyotrophic Lateral Sclerosis in Finland; phs000336, a Genome-Wide Association Study of Lung Cancer Risk; phs000346, Genome-Wide Association Study for Bladder Cancer Risk; phs000789, Collaborative Study of Genes, Nutrients and Metabolites (CSGNM); phs000206, Whole Genome Scan for Pancreatic Cancer Risk in the Pancreatic Cancer Cohort Consortium and Pancreatic Cancer Case–Control Consortium (PanScan); phs000297, eMERGE Network Study of the Genetic Determinants of Resistant Hypertension; phs000652, Cohort-Based Genome-Wide Association Study of Glioma (GliomaScan); phs000869, Barrett’s and Esophageal Adenocarcinoma Genetic Susceptibility Study (BEAGESS); phs000812, the Breast and Prostate Cancer Cohort Consortium (BPC3) GWAS of Aggressive Prostate Cancer and ER^−^ Breast Cancer; phs000428, Genetics Resource with the HRS; phs000360.v3, eMERGE Network Genome-Wide Association Study of Red Cell Indices, White Blood Count (WBC) Differential, Diabetic Retinopathy, Height, Serum Lipid Levels, Specifically Total Cholesterol, HDL (High Density Lipoprotein), LDL (Low Density Lipoprotein), and Triglycerides, and Autoimmune Hypothyroidism; phs000893.v1, Genome-Wide Association Study of Endometrial Cancer in the Epidemiology of Endometrial Cancer Consortium (E2C2); phs000168.v2, National Institute on Aging—Late Onset Alzheimer’s Disease Family Study: Genome-Wide Association Study for Susceptibility Loci; phs000092.v1, Study of Addiction: Genetics and Environment (SAGE); phs000864.v1, Genomic Predictors of Combat Stress Vulnerability and Resilience; phs000170.v2, a Genome-Wide Association Study on Cataract and HDL in the Personalized Medicine Research Project Cohort; phs000431.v2, IgA Nephropathy GWAS on Individuals of European Ancestry (IGANGWAS2); phs000237.v1, Northwestern NUgene Project: Type 2 Diabetes; phs000169.v1, Whole Genome Association Study of Visceral Adiposity in the Health Aging and Body Composition (Health ABC) Study; phs000982.v1, Genetic Analysis of Psoriasis and Psoriatic Arthritis: GWAS of Psoriatic Arthritis; phs000289.v2, National Human Genome Research Institute (NHGRI) GENEVA Genome-Wide Association Study of Venous Thrombosis (GWAS of VTE); phs000634.v1, National Cancer Institute (NCI) Genome Wide Association Study (GWAS) of Lung Cancer in Never Smokers; phs000274.v1, Genome-Wide Association Study of Celiac Disease; phs001172.v1, National Institute of Neurological Disorders and Stroke (NINDS) Parkinson’s Disease; phs000389.v1, GEnetics of Nephropathy—an International Effort (GENIE) GWAS of Diabetic Nephropathy in the UK GoKinD and All-Ireland Cohorts; phs000460.v1, Genetics of 24 Hour Urine Composition; phs000138.v2, GWAS for Genetic Determinants of Bone Fragility in European–American Premenopausal Women; phs000394.v1, Autopsy-Confirmed Parkinson Disease GWAS Consortium (APDGC); phs000948.v1, Genetic Discovery and Application in a Clinical Setting: Continuing a Partnership (eMERGE Phase II); phs000630.v1, Exome Chip Study of NIMH Controls; phs000678.v1, a Family-Based Study of Genes and Environment in Young-Onset Breast Cancer; phs000351.v1, National Cancer Institute Genome-Wide Association Study of Renal Cell Carcinoma; phs000314.v1, Genetic Associations in Idiopathic Talipes Equinovarus (Clubfoot)— GAIT; phs000147.v3, Cancer Genetic Markers of Susceptibility (CGEMS) Breast Cancer Genome-wide Association Study (GWAS)—Primary Scan: Nurses’ Health Study—Additional Cases: Nurses’ Health Study 2; phs000882.v1, National Cancer Institute (NCI) Prostate Cancer Genome-Wide Association Study for Uncommon Susceptibility Loci (PEGASUS); phs000238.v1, National Eye Institute Glaucoma Human Genetics Collaboration (NEIGHBOR) Consortium Glaucoma Genome-Wide Association Study; phs000397.v1, National Institute on Aging (NIA) Long Life Family Study (LLFS); phs000421.v1, a Genome-Wide Association Study of Fuchs’ Endothelial Corneal Dystrophy (FECD); phs000142.v1, a Whole Genome Association Scan for Myopia and Glaucoma Endophenotypes using Twin Studies; phs000303.v1, Genetic Epidemiology of Refractive Error in the KORA (Kooperative Gesundheitsforschung in der Region Augsburg) Study; phs000125.v1, CIDR: Collaborative Study on the Genetics of Alcoholism Case Control Study; phs001039.v1, International Age-Related Macular Degeneration Genomics Consortium—Exome Chip Experiment; phs000187.v1, High Density SNP Association Analysis of Melanoma: Case–Control and Outcomes Investigation; phs000101.v5, Genome-Wide Association Study of Amyotrophic Lateral Sclerosis; phs002068.v1.p1, Sporadic ALS Australia Systems Genomics Consortium (SALSA-SGC)). Gene expression data used in the preparation of this article were obtained from the ANSWER ALS Data Portal (AALS-01184). For up-to-date information on the study, visit https://dataportal.answerals.org.

## References

1. Ryan, M., Heverin, M., McLaughlin, R. L. & Hardiman, O. Lifetime Risk and Heritability of Amyotrophic Lateral Sclerosis. JAMA Neurol. 76, 1367–1374 (2019).

2. van Rheenen, W. et al. Common and rare variant association analyses in amyotrophic lateral sclerosis identify 15 risk loci with distinct genetic architectures and neuron-specific biology. Nat. Genet. 53, 1636–1648 (2021).

3. Bulik-Sullivan, B. K. et al. LD Score regression distinguishes confounding from polygenicity in genome-wide association studies. Nat. Genet. 47, 291–295 (2015).

4. Bulik-Sullivan, B. et al. An atlas of genetic correlations across human diseases and traits. Nat. Genet. 47, 1236–1241 (2015).

5. Xiao, L. et al. Multiple-Tissue Integrative Transcriptome-Wide Association Studies Discovered New Genes Associated With Amyotrophic Lateral Sclerosis. Front. Genet. 11, 587243 (2020).

6. Baxi, E. G. et al. Answer ALS, a large-scale resource for sporadic and familial ALS combining clinical and multiomics data from induced pluripotent cell lines. Nat. Neurosci. 25, 226–237 (2022).

7. Finucane, H. K. et al. Partitioning heritability by functional annotation using genome-wide association summary statistics. Nat. Genet. 47, 1228–1235 (2015).

8. Harrington, A. J. et al. MEF2C regulates cortical inhibitory and excitatory synapses and behaviors relevant to neurodevelopmental disorders. eLife 5, e20059 (2016).

9. Cosgrove, D. et al. Genes influenced by MEF2C contribute to neurodevelopmental disease via gene expression changes that affect multiple types of cortical excitatory neurons. Hum. Mol. Genet. 30, 961–970 (2021).

10. Manjaly, Z. R. et al. The sex ratio in amyotrophic lateral sclerosis: A population based study. Amyotroph. Lateral Scler. Off. Publ. World Fed. Neurol. Res. Group Mot. Neuron Dis. 11, 439–442 (2010).

11. Arosio, A. et al. MEF2D and MEF2C pathways disruption in sporadic and familial ALS patients. Mol. Cell. Neurosci. 74, 10–17 (2016).

12. Darlington, D., Schiller, M., Mains, R. & Eipper, B. The expression of regulated endocrine-specific protein of 18 kDa in peptidergic cells of rat peripheral endocrine tissues and in blood. J. Endocrinol. 155, 329–341 (1997).

13. Su, J. et al. RESP18 deficiency has protective effects in dopaminergic neurons in an MPTP mouse model of Parkinson’s disease. Neurochem. Int. 118, 195–204 (2018).

14. Bellenguez, C. et al. New insights into the genetic etiology of Alzheimer’s disease and related dementias. Nat. Genet. 54, 412–436 (2022).

15. Nagle, M. W. et al. The 4p16.3 Parkinson Disease Risk Locus Is Associated with GAK Expression and Genes Involved with the Synaptic Vesicle Membrane. PLOS ONE 11, e0160925 (2016).

16. Li, Y. et al. Identification of novel genes for age-at-onset of Alzheimer’s disease by combining quantitative and survival trait analyses. Alzheimers Dement. 19, 3148–3157 (2023).

17. Camu, W. et al. Repeated 5-day cycles of low dose aldesleukin in amyotrophic lateral sclerosis (IMODALS): A phase 2a randomised, double-blind, placebo-controlled trial. EBioMedicine 59, 102844 (2020).

18. Falconer, D. S. The inheritance of liability to diseases with variable age of onset, with particular reference to diabetes mellitus. Ann. Hum. Genet. 31, 1–20 (1967).

19. Zhou, W. et al. Efficiently controlling for case-control imbalance and sample relatedness in large-scale genetic association studies. Nat. Genet. 50, 1335–1341 (2018).

20. Willer, C. J., Li, Y. & Abecasis, G. R. METAL: fast and efficient meta-analysis of genomewide association scans. Bioinforma. Oxf. Engl. 26, 2190–2191 (2010).

21. Grotzinger, A. D., Fuente, J. de la, Privé, F., Nivard, M. G. & Tucker-Drob, E. M. Pervasive Downward Bias in Estimates of Liability-Scale Heritability in Genome-wide Association Study Meta-analysis: A Simple Solution. Biol. Psychiatry 93, 29–36 (2023).

22. van Rheenen, W. et al. Genome-wide association analyses identify new risk variants and the genetic architecture of amyotrophic lateral sclerosis. Nat. Genet. 48, 1043–1048 (2016).

23. Gao, F. et al. XWAS: A Software Toolset for Genetic Data Analysis and Association Studies of the X Chromosome. J. Hered. 106, 666–671 (2015).

24. Gusev, A. et al. Integrative approaches for large-scale transcriptome-wide association studies. Nat. Genet. 48, 245–252 (2016).

