## Supplementary_figs_tabs_text for "Sex-specific risk loci and modified *MEF2C* expression in ALS"

### Supplementary material

#### Supplementary table 1 Sample sizes for strata used in sex-specific GWAS analysis

| | **Male** | | **Female** | |
|---|---|---|---|---|
| **Stratum** | **ALS** | **Control** | **ALS** | **Control** |
| **s1** | 1,358 | 4,864 | 890 | 6,264 |
| **s2** | 864 | 1,434 | 594 | 609 |
| **s3** | 970 | 1,118 | 726 | 1,430 |
| **s4** | 2,064 | 17,967 | 1,328 | 23,278 |
| **s6** | 8,357 | 15,652 | 5,951 | 14,908 |
| **s7** | 1,934 | 9,110 | 1,406 | 10,573 |
| **Total** | **15,547** | **50,145** | **10,895** | **57,062** |

#### Supplementary table 2 Significant loci in sex-specific GWAS

| **Significant in female-specific GWAS** | | | | | | | | **Closest** | **Sex-** |
|---|---|---|---|---|---|---|---|---|---|
| **Locus*** | **Lead SNP** | **A1** | **A2** | **$ß_{female}$ (SE)** | **$ß_{male}$ (SE)** | **$p_{female}$** | **$p_{male}$** | **gene** | **specific†** |
| chr4:994011-1020391 | rs74921869 | A | G | 0.124 (0.023) | -0.004 (0.021) | $9.34 \times 10^{-8}$ | 0.85 | *FGFRL1* | Yes |
| chr5:88102863-88144520 | rs304152 | T | G | 0.100 (0.019) | 0.024 (0.016) | $5.46 \times 10^{-8}$ | 0.14 | *MEF2C* | Yes |
| chr9:26572913-28669828 | rs2453555 | A | G | 0.193 (0.020) | 0.155 (0.017) | $5.49 \times 10^{-23}$ | $1.73 \times 10^{-19}$ | *C9orf72* | No |
| chr19:17742469-17755139 | rs12608932 | A | C | -0.176 (0.019) | -0.101 (0.017) | $6.84 \times 10^{-21}$ | $1.25 \times 10^{-9}$ | *UNC13A* | No |
| chr21:32938642-33128270 | rs80265967 | A | C | -1.108 (0.177) | -1.022 (0.193) | $3.45 \times 10^{-10}$ | $1.11 \times 10^{-7}$ | *SOD1* | No |
| **Significant in male-specific GWAS** | | | | | | | | **Closest** | **Sex-** |
| **Locus*** | **Lead SNP** | **A1** | **A2** | **$ß_{female}$** | **$ß_{male}$** | **$p_{female}$** | **$p_{male}$** | **gene** | **specific†** |
| chr2:56611289-57878262 | rs80178959 | T | C | -0.228 (0.220) | -1.012 (0.191) | 0.30 | $1.17 \times 10^{-7}$ | *CCDC85A* | Yes |
| chr2:220151858-220209246 | rs6738336 | C | G | 0.036 (0.019) | 0.088 (0.017) | 0.06 | $1.30 \times 10^{-7}$ | *RESP18* | Yes |
| chr6:32552072-32682429 | rs9275477 | A | C | 0.120 (0.030) | 0.153 (0.026) | $4.9 \times 10^{-5}$ | $4.86 \times 10^{-9}$ | *HLA* | No |
| chr7:5202904-5202904 | rs13241583 | A | G | 0.005 (0.042) | -0.199 (0.037) | 0.90 | $7.19 \times 10^{-8}$ | *WIPI2* | Yes |
| chr9:27469573-27597891 | rs2484319 | A | C | -0.190 (0.020) | -0.156 (0.017) | $1.79 \times 10^{-22}$ | $1.11 \times 10^{-19}$ | *C9orf72* | No |
| chr11:24240182-24409277 | rs80027360 | T | C | 0.097 (0.070) | -0.325 (0.062) | 0.17 | $1.55 \times 10^{-7}$ | *LUZP2* | Yes |
| chr14:31000949-31204433 | rs7152417 | T | C | -0.077 (0.018) | -0.097 (0.016) | $1.95 \times 10^{-5}$ | $7.29 \times 10^{-10}$ | *SCFD1* | No |
| chr19:17742469-17753239 | rs12973192 | C | G | -0.176 (0.019) | -0.101 (0.017) | $9.48 \times 10^{-21}$ | $1.22 \times 10^{-9}$ | *UNC13A* | No |
| chr21:32938642-33128270 | rs80265967 | A | C | -1.108 (0.177) | -1.022 (0.193) | $3.45 \times 10^{-10}$ | $1.11 \times 10^{-7}$ | *SOD1* | No |

Effect sizes are reported for allele 1 (A1)
† Defined as $p_{sex1} < 2.3 \times 10^{-7}$ and $p_{sex2} > 0.05$
* Locus boundaries defined by FUMA

#### Supplementary table 3 Significant locus in X-chromosome analysis

| **Locus** | **Lead SNP** | **A1** | **A2** | **$ß_{female}$ (SE)** | **$ß_{male}$ (SE)** | **$p_{female}$** | **$p_{male}$** | **$p_{combined}$** | **Closest gene** |
|---|---|---|---|---|---|---|---|---|---|
| chrX:104409884 | rs17332232 | T | C | -0.197 (0.039) | -0.150 (0.048) | $7.84 \times 10^{-7}$ | $2.51 \times 10^{-3}$ | $4.14 \times 10^{-8}$ | *IL1RAPL2* |

Effect sizes are reported for allele 1 (A1)

#### Supplementary table 4 Sex-specific significant loci in sex-specific TWAS

| **Gene** | **$p_{female}$** | **$p_{male}$** |
|---|---|---|
| *RESP18* | 0.12 | $5.22 \times 10^{-7}$ |
| *MEF2C* | $1.26 \times 10^{-7}$ | 0.13 |

**Supplementary table 5 Partitioned heritability estimates for sex-stratified GWAS statistics human orthologues of differentially expressed genes in *Mef2c*-conditional knockout (cKO) mice.**

| Direction in *Mef2c* cKO | GWAS | Enrichment | SE | p |
|---|---|---|---|---|
| All | Both sexes | 1.73 | 0.30 | 0.0112 |
| Upregulated | Both sexes | 1.81 | 0.58 | 0.1611 |
| Downregulated | Both sexes | 1.74 | 0.31 | 0.0130 |
| All | Female | 1.66 | 0.42 | 0.0945 |
| Upregulated | Female | 0.58 | 0.76 | 0.5633 |
| Downregulated | Female | 2.40 | 0.62 | 0.0026* |
| All | Male | 0.32 | 1.52 | 0.5898 |
| Upregulated | Male | 1.47 | 2.28 | 0.8274 |
| Downregulated | Male | -0.17 | 2.08 | 0.4673 |

GWAS of both sexes is the European subset of the 2021 ALS GWAS[2].
* $p < 0.006$ (corrected for 9 tests)

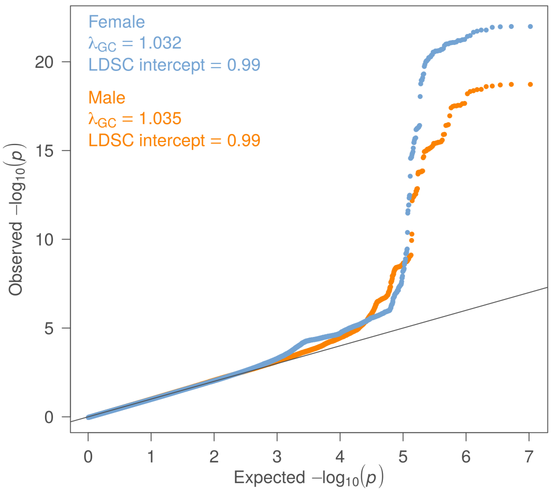


**Supplementary figure 1 Quantile-quantile plot for female-specific and male-specific GWAS statistics.**

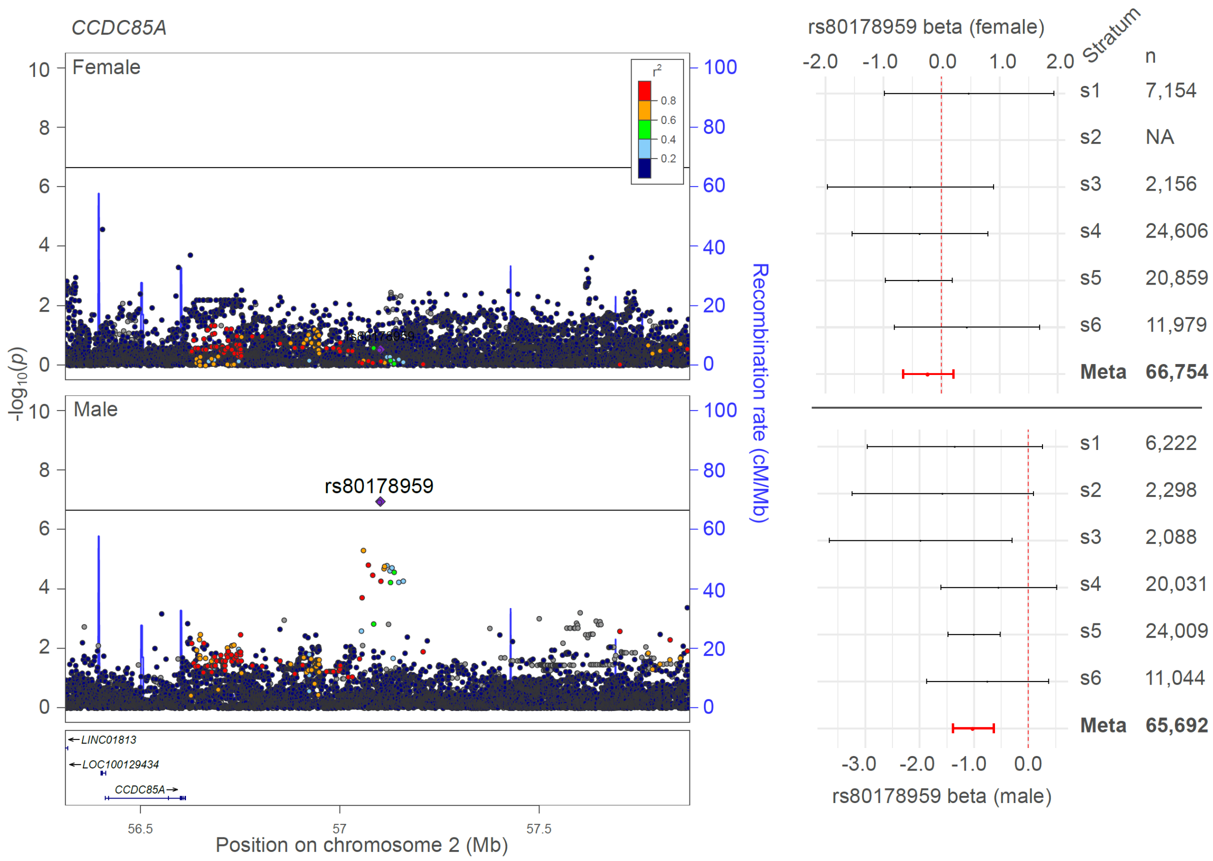


**Supplementary figure 2 Locus- and cohort-level detail for rs80178959 (*CCDC85A*)** P-value threshold corresponds to our 1% FDR cutoff ($p<2.3\times10^{-7}$)

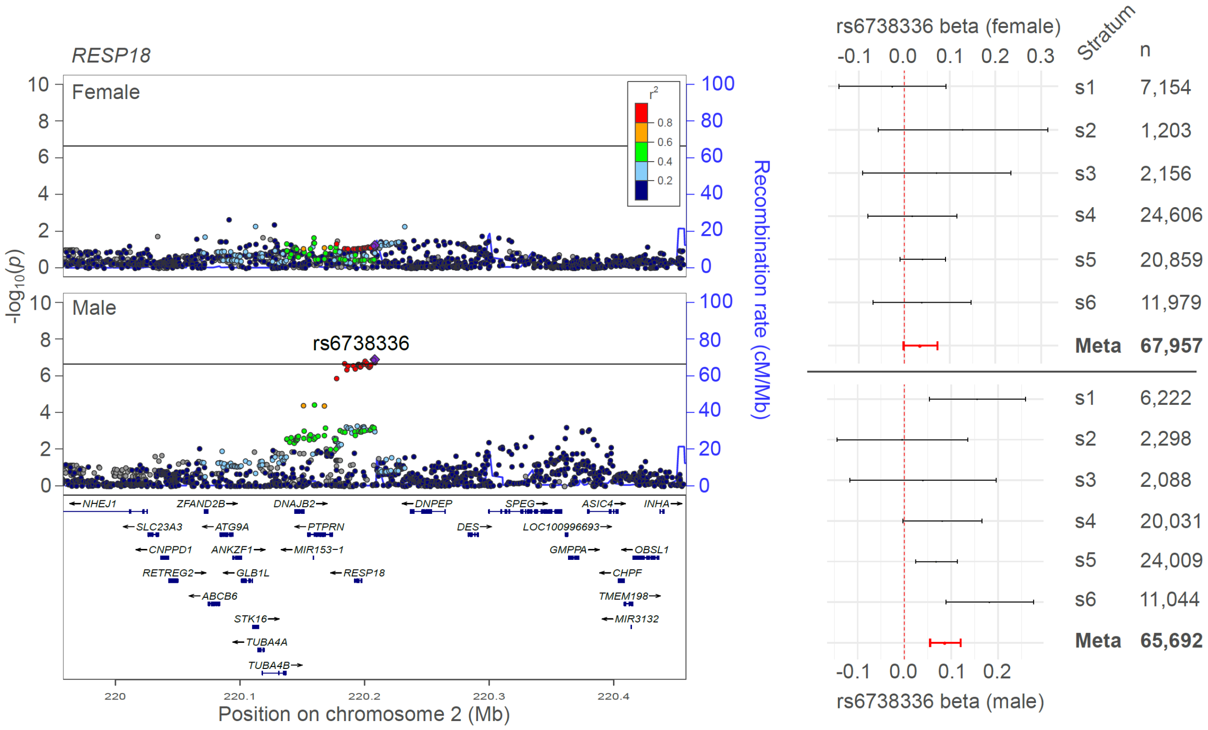


**Supplementary figure 3 Locus- and cohort-level detail for rs6738336 (*RESP18*)** P-value threshold corresponds to our 1% FDR cutoff ($p<2.3\times10^{-7}$)

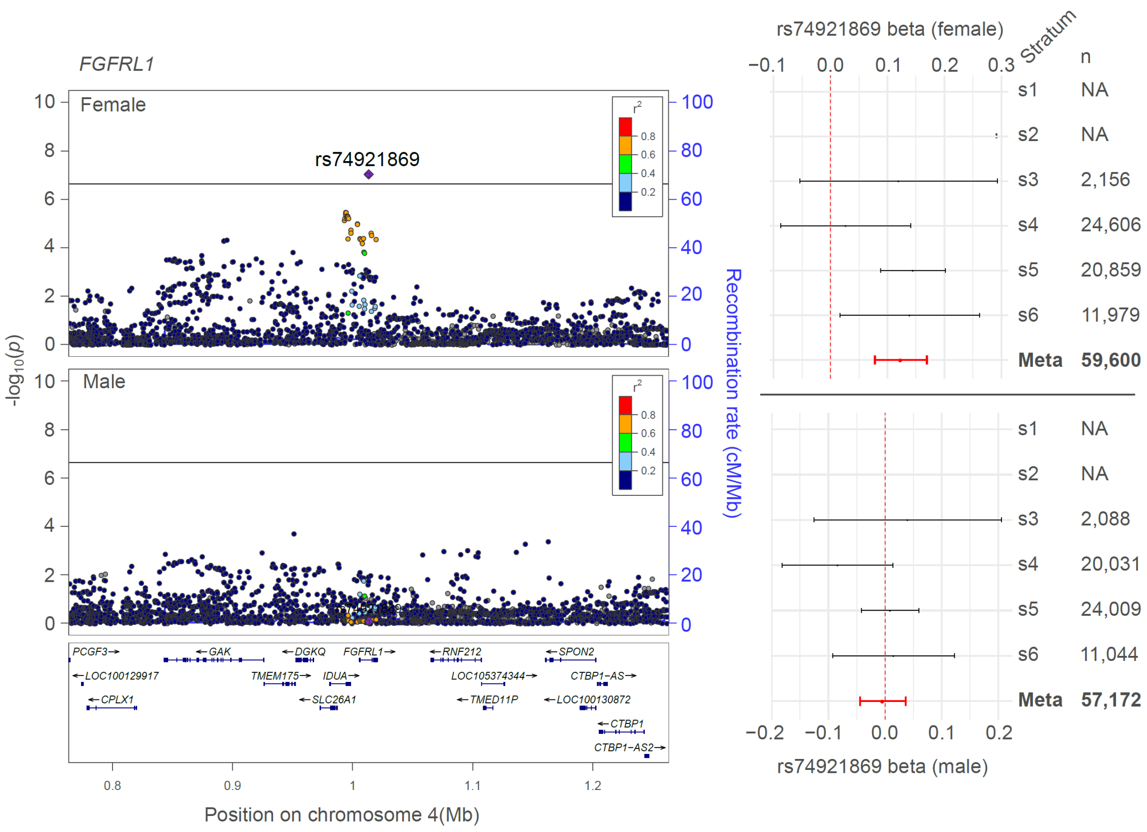


**Supplementary figure 4 Locus- and cohort-level detail for rs74821869 (*FGFLR1*)** P-value threshold corresponds to our 1% FDR cutoff ($p<2.3\times10^{-7}$)

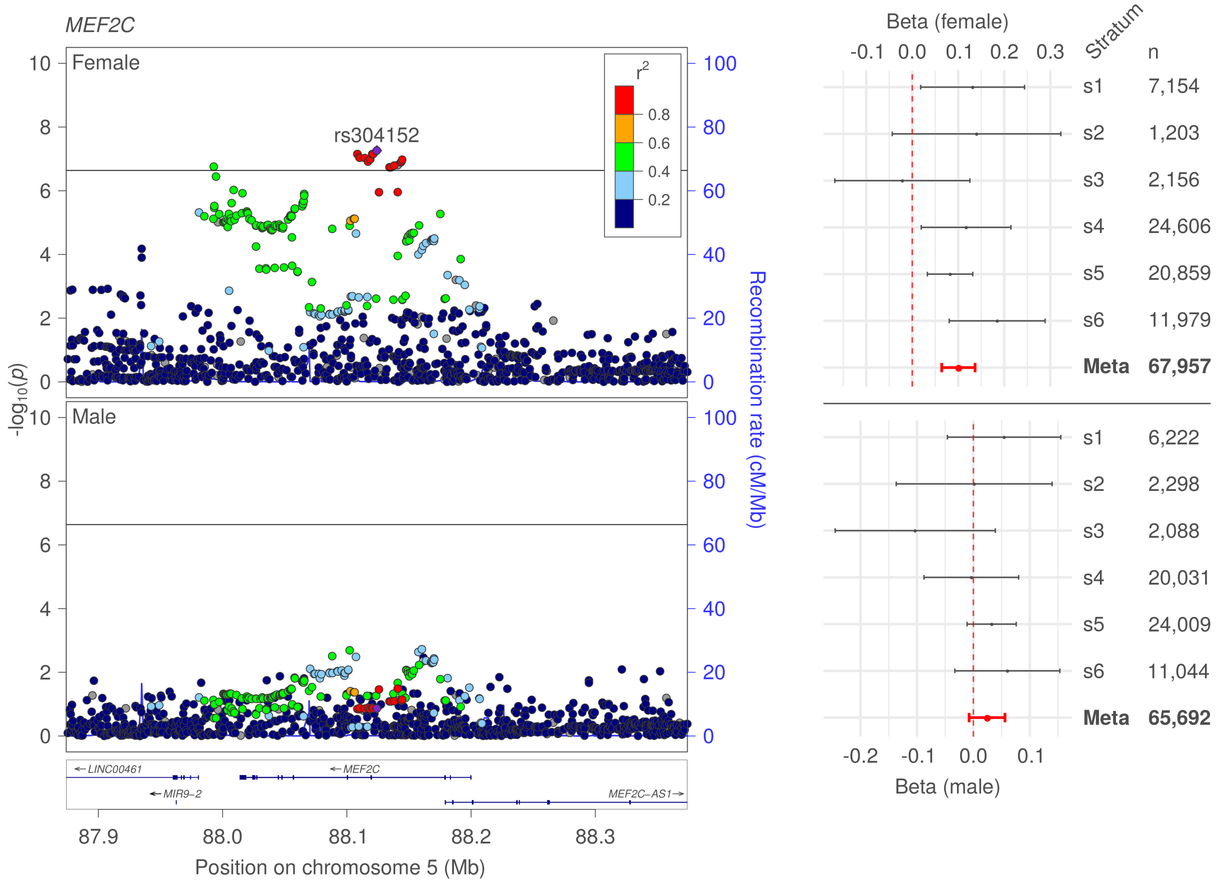


**Supplementary figure 5 Locus- and cohort-level detail for rs304152 (*MEF2C*)** P-value threshold corresponds to our 1% FDR cutoff ($p<2.3\times10^{-7}$)

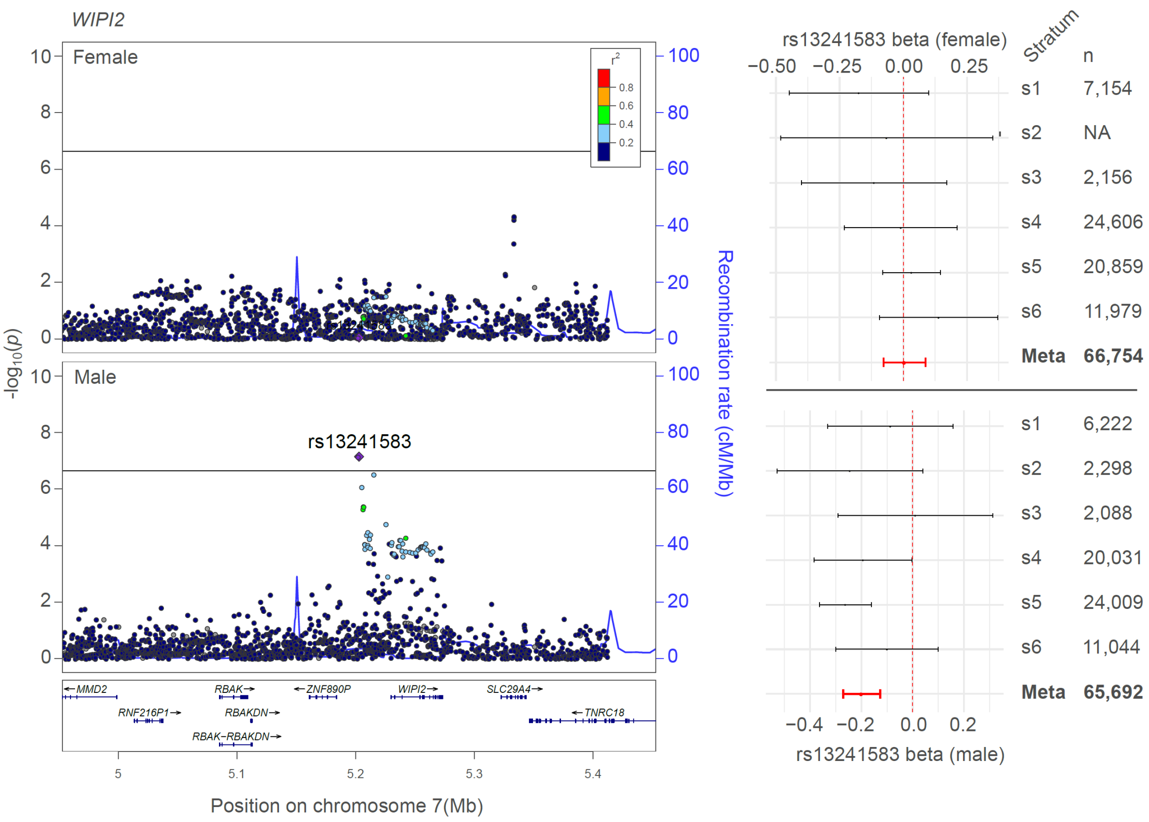


**Supplementary figure 6 Locus- and cohort-level detail for rs13241583 (*WIPI2*)** P-value threshold corresponds to our 1% FDR cutoff ($p<2.3\times10^{-7}$)

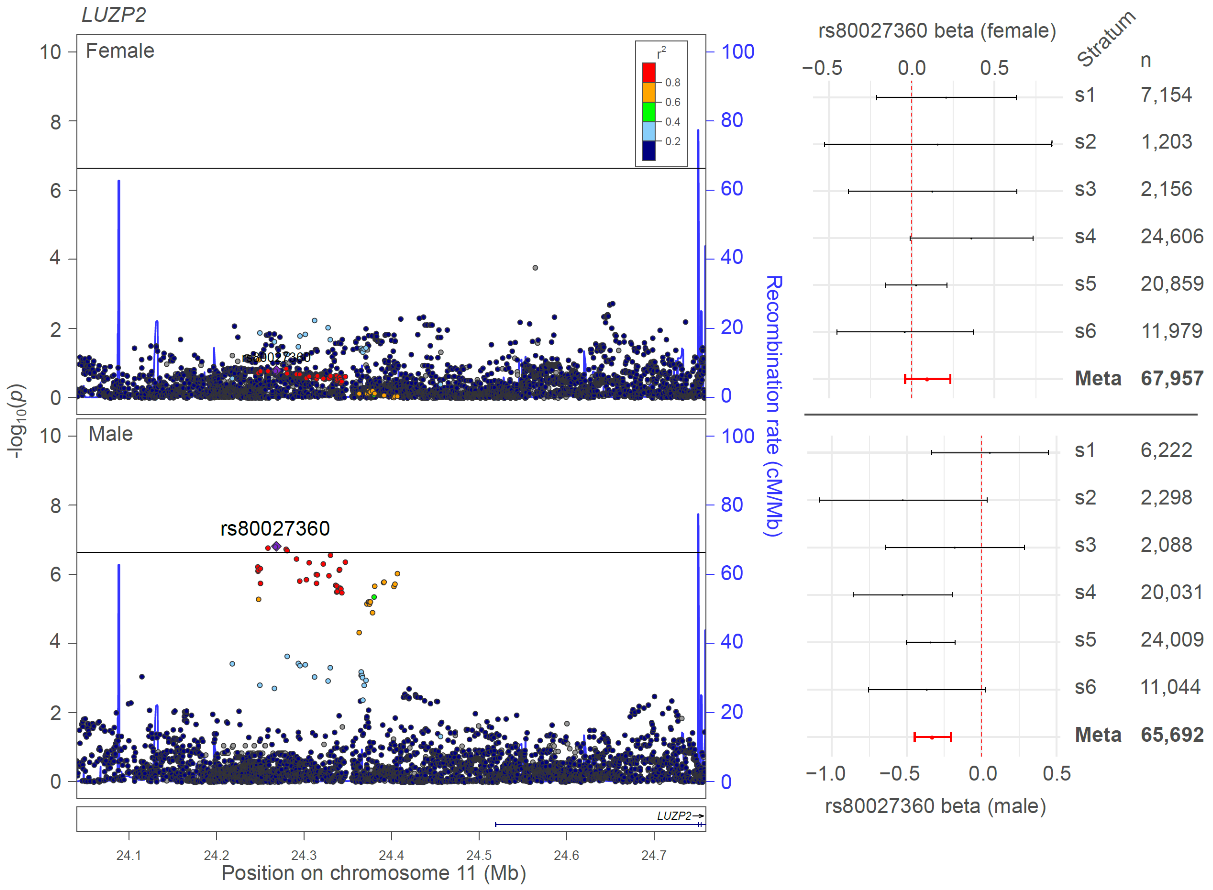


**Supplementary figure 7 Locus- and cohort-level detail for rs80027360 (*LUZP2*)** P-value threshold corresponds to our 1% FDR cutoff ($p<2.3\times10^{-7}$)

### Supplementary note 1

Project MinE ALS GWAS Consortium Authors

Wouter van Rheenen[1], Mark K. Bakker[1], Kristel R. van Eijk[1], Maarten Kooyman[1], Ross P. Byrne[2], Ahmad Al Khleifat[3,4], Alfredo Iacoangeli[3,4,5,6], Nicola Ticozzi[7,8], Johnathan Cooper-Knock[9], Marta Gromicho[10], Siddharthan Chandran[11,12], Karen E. Morrison[13], Pamela J. Shaw[9], John Hardy[14], Michael Sendtner[15], Thomas Meyer[16], Nazli Başak[17], Isabella Fogh[3], Adriano Chiò[18,19], Andrea Calvo[18,19], Elisabetta Pupillo[20], Giancarlo Logroscino[21], Marc Gotkine[22,23], Yossef Lerner[22,23], Patrick Vourc'h[24,25], Philippe Corcia[25,26], Philippe Couratier[27,28], Stéphanie Millecamps[29], François Salachas[29,30], Gaëlle Bruneteau[29,30], Jesus S. Mora Pardina[31], Ricardo Rojas-García[32], Patrick Dion[33], Jay P. Ross[33,34], Albert C. Ludolph[35], Jochen H. Weishaupt[36], Axel Freischmidt[35], Gilbert Bensimon[37,38,39,40], Lukas Tittmann[41], Wolfgang Lieb[41], Andre Franke[42], Stephan Ripke[43,44,45], David C. Whiteman[46], Catherine M. Olsen[46], Andre G. Uitterlinden[47,48], Clifton L. Dalgard[49,50], Albert Hofman[47], Philippe Amouyel[51], Andrew B. Singleton[52], Miguel Mitne Neto[53], Ruben J. Cauchi[54], Roel A. Ophoff[55,56,57], Vivianna M. Van Deerlin[58], Julian Grosskreutz[59,60], Caroline Graff[61], Lev Brylev[62,63,64], Vera Fominikh[62], Boris Rogelj[65,66], Blaž Koritnik[67], Janez Zidar[67], Zorica Stević[68], Vivian Drory[69,70], Monica Povedano[71], Ian P. Blair[72], Matthew C. Kiernan[73], Garth A. Nicholson[72,74,75], Anjali K. Henders[76], Mamede de Carvalho[10], Susana Pinto[10], Susanne Petri[77], Markus Weber[78], Guy A. Rouleau[79], Vincenzo Silani[7,8], Jonathan Glass[80], Robert H. Brown[81], John E. Landers[81], Christopher E. Shaw[3], Peter M. Andersen[82], Fleur C. Garton[76], Allan F. McRae[76], Russell L. McLaughlin[2], Orla Hardiman[83], Kevin P. Kenna[1,84], Ammar Al-Chalabi[3,85], Philip Van Damme[86,87], Leonard H. van den Berg[1], Jan H. Veldink[1]

1. Department of Neurology, UMC Utrecht Brain Center, University Medical Center Utrecht, Utrecht University, Utrecht, The Netherlands.
2. Complex Trait Genomics Laboratory, Smurfit Institute of Genetics, Trinity College Dublin, Dublin D02 PN40, Ireland.
3. Maurice Wohl Clinical Neuroscience Institute, Department of Basic and Clinical Neuroscience, Institute of Psychiatry, Psychology & Neuroscience, King's College London, London, UK.
4. Perron Institute for Neurological and Translational Science, University of Western Australia Medical School, Perth, WA 6009, Australia
5. Department of Biostatistics and Health Informatics, Institute of Psychiatry, Psychology and Neuroscience, King's College London, London, UK.
6. National Institute for Health Research Biomedical Research Centre and Dementia Unit, South London and Maudsley NHS Foundation Trust and King's College London, London, UK.
7. Department of Neurology-Stroke Unit and Laboratory of Neuroscience, Istituto Auxologico Italiano IRCCS, Milan, Italy.
8. Department of Pathophysiology and Transplantation, "Dino Ferrari" Center, Università degli Studi di Milano, Milan, Italy.
9. Sheffield Institute for Translational Neuroscience (SiTraN), School of Medicine and Population Health, University of Sheffield, Sheffield, UK
10. Instituto de Fisiologia, Instituto de Medicina Molecular João Lobo Antunes, Faculdade de Medicina, Universidade de Lisboa, Lisbon, Portugal.
11. Euan MacDonald Centre for Motor Neurone Disease Research, Edinburgh, UK.

12. Centre for Neuroregeneration and Medical Research Council Centre for Regenerative Medicine, University of Edinburgh, Edinburgh, UK.
13. School of Medicine, Dentistry, and Biomedical Sciences, Queen's University Belfast, Belfast, UK.
14. Department of Molecular Neuroscience, Institute of Neurology, University College London, London, UK.
15. Institute of Clinical Neurobiology, University Hospital Würzburg, Würzburg, Germany.
16. Charité University Hospital, Humboldt-University, Berlin, Germany.
17. Neurodegeneration Research Laboratory, Bogazici University, Istanbul, Turkey.
18. "Rita Levi Montalcini" Department of Neuroscience, ALS Centre, University of Torino, Turin, Italy.
19. Neurologia 1, Azienda Ospedaliero Universitaria Città della Salute e della Scienza, Turin, Italy.
20. Research Center for ALS, (Department of Neuroscience, Laboratory of Neurological Diseases), Istituto di Ricerche Farmacologiche Mario Negri IRCCS, Milan, Italy
21. Department of Clinical Research in Neurology, University of Bari at "Pia Fondazione Card G. Panico" Hospital, Bari, Italy.
22. Faculty of Medicine, Hebrew University of Jerusalem, Jerusalem, Israel.
23. Department of Neurology, Agnes Ginges Center for Human Neurogenetics, Hadassah Medical Organization, Jerusalem, Israel.
24. Service de Biochimie et Biologie moléculaire, CHU de Tours, Tours, France.
25. UMR 1253, Université de Tours, Inserm, 37044 Tours, France.
26. Centre de référence sur la SLA, CHU de Tours, Tours, France.
27. Centre de référence sur la SLA, CHRU de Limoges, Limoges, France.
28. UMR 1094, Université de Limoges, Inserm, 87025 Limoges, France.
29. Sorbonne Université, Institut du Cerveau - Paris Brain Institute - ICM, Inserm, CNRS, APHP, Hôpital de la Pitié Salpêtrière, Paris, France
30. Département de Neurologie, Centre de référence SLA Ile de France, Hôpital de la Pitié Salpêtrière, AP-HP, Paris, France.
31. ALS Unit, Hospital Universitario San Rafael, Madrid, Spain
32. MND Clinic, Neurology Department, Hospital de la Santa Creu i Sant Pau de Barcelona, Universitat Autonoma de Barcelona, Barcelona, Spain.
33. The Montréal Neurological Institute-Hospital, McGill University, Montréal, QC H3A 2B4, Canada.
34. Department of Human Genetics, McGill University, Montréal, QC H3A 0C7, Canada.
35. Department of Neurology, Ulm University, Ulm, Germany.
36. Division of Neurodegeneration, Department of Neurology, University Medicine Mannheim, Medical Faculty Mannheim, Heidelberg University, Mannheim, Germany.
37. Département de Pharmacologie Clinique, Hôpital de la Pitié-Salpêtrière, UPMC Pharmacologie, AP-HP, Paris, France.
38. Pharmacologie Sorbonne Université, Paris, France.
39. Institut du Cerveau, Paris Brain Institute ICM, Paris, France.
40. Laboratoire de Biostatistique, Epidémiologie clinique, Santé Publique Innovation et Méthodologie (BESPIM), CHU-Nîmes, Nîmes, France.
41. Popgen Biobank and Institute of Epidemiology, Christian Albrechts-University Kiel, Kiel, Germany.

42. Institute of Clinical Molecular Biology, Kiel University, Kiel, Germany.
43. Analytic and Translational Genetics Unit, Massachusetts General Hospital, Boston, Massachusetts, USA.
44. Stanley Center for Psychiatric Research, Broad Institute of MIT and Harvard, Cambridge, Massachusetts, USA.
45. Department of Psychiatry and Psychotherapy, Charité - Universitätsmedizin, Berlin 10117, Germany.
46. Cancer Control Group, QIMR Berghofer Medical Research Institute, Herston, QLD, Australia.
47. Department of Internal Medicine, Genetics Laboratory, Erasmus Medical Center Rotterdam, Rotterdam, The Netherlands.
48. Department of Epidemiology, Erasmus Medical Center Rotterdam, Rotterdam, The Netherlands.
49. Department of Anatomy, Physiology and Genetics, Uniformed Services University of the Health Sciences, Bethesda, MD, 20814, USA
50. The American Genome Center, Center for Military Precision Health, Uniformed Services University of the Health Sciences, Bethesda, MD, USA
51. Univ. Lille, Inserm, Centre Hosp. Univ. Lille, Institut Pasteur de Lille, UMR1167 - RID-AGE LabEx DISTALZ - Risk factors and molecular determinants of aging-related diseases, F-59000 Lille, France.
52. Molecular Genetics Section, Laboratory of Neurogenetics, National Institute on Aging, NIH, Porter Neuroscience Research Center, Bethesda, Maryland, USA.
53. Universidade de São Paulo, São Paulo, Brazil.
54. Centre for Molecular Medicine and Biobanking & Department of Physiology and Biochemistry, Faculty of Medicine and Surgery, University of Malta, Malta.
55. University Medical Center Utrecht, Department of Psychiatry, Rudolf Magnus Institute of Neuroscience, The Netherlands
56. Department of Human Genetics, David Geffen School of Medicine, University of California, Los Angeles, California, USA.
57. Center for Neurobehavioral Genetics, Semel Institute for Neuroscience and Human Behavior, University of California, Los Angeles, California, USA.
58. Center for Neurodegenerative Disease Research, Department of Pathology and Laboratory Medicine, Perelman School of Medicine at the University of Pennsylvania, Philadelphia, Pennsylvania, USA.
59. Hans-Berger-Department of Neurology, Jena University Hospital, Jena, Germany.
60. Precision Neurology Unit, Department of Neurology, University Hospital Schleswig-Holstein, University of Luebeck, Luebeck, Germany.
61. Department of Geriatric Medicine, Karolinska University Hospital-Huddinge, Stockholm, Sweden.
62. Department of Neurology, Bujanov Moscow Clinical Hospital, Moscow, Russia.
63. Moscow Research and Clinical Center for Neuropsychiatry of the Healthcare Department, Moscow, Russia.
64. Department of Functional Biochemistry of the Nervous System, Institute of Higher Nervous Activity and Neurophysiology Russian Academy of Sciences, Moscow, Russia.
65. Department of Biotechnology, Jožef Stefan Institute, Ljubljana, Slovenia.
66. Faculty of Chemistry and Chemical Technology, Universtiy of Ljubljana, Ljubljana, Slovenia.
67. Ljubljana ALS Centre, Institute of Clinical Neurophysiology, University Medical Centre Ljubljana, Ljubljana, Slovenia.

68. Clinic of Neurology, Clinical Center of Serbia, School of Medicine, University of Belgrade, Belgrade, Serbia.
69. Neuromuscular Diseases Unit, Department of Neurology, Tel Aviv Sourasky Medical Center, Tel Aviv, Israel.
70. Faculty of Medicine, Tel Aviv University, Tel Aviv, Israel.
71. Functional Unit of Amyotrophic Lateral Sclerosis (UFELA), Service of Neurology, Bellvitge University Hospital, L'Hospitalet de Llobregat, Barcelona, Spain.
72. Centre for Motor Neuron Disease Research, Faculty of Medicine, Health and Human Sciences, Macquarie University, NSW 2109, Australia.
73. Brain and Mind Centre, University of Sydney, Sydney, New South Wales, Australia.
74. Northcott Neuroscience Laboratory, ANZAC Research Institute, Concord, NSW 2139, Australia.
75. Molecular Medicine Laboratory, Concord Repatriation General Hospital, Concord, NSW 2139, Australia.
76. Institute for Molecular Bioscience, University of Queensland, Brisbane, Queensland, Australia.
77. Department of Neurology, Hannover Medical School, Hannover, Germany.
78. Neuromuscular Diseases Unit/ALS Clinic, Kantonsspital St. Gallen, 9007, St. Gallen, Switzerland.
79. Montreal Neurological Institute and Hospital, McGill University, Montréal H3A 2B4, Canada.
80. Department Neurology, Emory University School of Medicine, Atlanta, Georgia, USA.
81. Department of Neurology, University of Massachusetts Medical School, Worcester, Massachusetts, USA.
82. Department of Clinical Sciences, Neurosciences, Umeå University, SE-901 85 Umeå, Sweden.
83. Academic Unit of Neurology, Trinity Biomedical Sciences Institute, Trinity College Dublin, Dublin D02 PN40, Ireland.
84. Department of Translational Neuroscience, UMC Utrecht Brain Center, University Medical Center Utrecht, Utrecht University, Utrecht, The Netherlands.
85. King's College Hospital, Denmark Hill, SE5 9RS London, UK.
86. KU Leuven – University of Leuven, Department of Neurosciences, Experimental Neurology, and Leuven Brain Institute (LBI), Leuven, Belgium.
87. University Hospitals Leuven, Department of Neurology, Leuven, Belgium.

### Supplementary note 2

International ALS/FTD Genomics Consortium

Robert H Baloh[1], Robert Bowser[2], Alexis Brice[3,4], James Broach[5], Roy H Campbell[6], Adriano Chiò[7,8], John Cooper-Knock[9], Daniele Cusi[10], Carsten Drepper[11], Vivian E Drory[12], Travis L Dunckley[13], Eva L Feldman[14], Pietro Fratta[15], Summer B Gibson[16], Jonathan D Glass[17], Stephen A Goutman[14], John Hardy[18], Matthew B Harms[19], Terry D Heiman-Patterson[20,21], Lilja Jansson[22], Janine Kirby[9], Neil W Kowall[23], Hannu Laaksovirta[22], John E Landers[24], Francesco Landi[25], Isabelle Le Ber[3,4], Claire Guissart[26], Daniel JL MacGowan[27], Nicholas J Maragakis[28], Kevin Mouzat[26], Liisa Myllykangas[29], Richard W Orrell[30], Erik P Pioro[31], Stefan M Pulst[16], John M Ravits[32], Ekaterina Rogaeva[33], Sara Rollinson[34], Jeffrey D Rothstein[28], Erika Salvi[10], Sonja W Scholz[35,28], Michael Sendtner[36], Pamela J Shaw[9], Katie C Sidle[18], Zachary Simmons[37], David J Stone[38], Pentti J Tienari[22], Bryan J Traynor[39,28], Juan C Troncoso[40], Miko Valori[22], Philip Van Damme[41,42], Vivianna M Van Deerlin[43], Ludo Van Den Bosch[41,44], Lorne Zinman[45]

1. Department of Neurology, Cedars-Sinai Medical Center, Los Angeles, CA, 90048, USA
2. Division of Neurology, Barrow Neurological Institute, Phoenix, AZ, 85013, USA
3. Centre de Recherche de l'Institut du Cerveau et de la Moelle épinière, Université Pierre et Marie Curie, Paris, France
4. INSERM U975, Paris, France
5. Department of Biochemistry, Penn State College of Medicine, Hershey, PA, 17033, USA
6. Department of Computer Science, University of Illinois at Urbana-Champaign, 201 North Goodwin Avenue, Urbana, IL, 61801, USA
7. 'Rita Levi Montalcini' Department of Neuroscience, University of Turin, Via Verdi 8, Turin, 101246, Italy
8. C.N.R., Via S. Martino della Battaglia, 44, Rome, 00185, Italy
9. Sheffield Institute for Translational Neuroscience (SiTraN), School of Medicine and Population Health, University of Sheffield, Sheffield, UK
10. Scientific Unit, Bio4Dreams - Business Nursery for Life Sciences, Milan, Italy.
11. Institute for Clinical Neurobiology, University of Würzburg, Würzburg, D-97078, Germany
12. Department of Neurology, Tel-Aviv Sourasky Medical Center, Tel-Aviv, Israel
13. Neurogenomics Division, Translational Genomics Research Institute, Phoenix, AZ, USA
14. Department of Neurology, University of Michigan, 1500 E Medical Center Dr, Ann Arbor, MI, 48109, USA
15. Sobell Department of Motor Neuroscience and Movement Disorders, Institute of Neurology, University College London, London, WC1N 3BG, UK
16. Department of Neurology, University of Utah School of Medicine, 175 North Medical Drive East, Salt Lake City, UT, 84132, USA
17. Department of Neurology, Emory University School of Medicine, Atlanta, GA, 30322, USA
18. Department of Molecular Neuroscience and Reta Lila Weston Laboratories, Institute of Neurology, University College London, London, WC1N 3BG, UK
19. Department of Neurology, Columbia University, New York, NY, 10032, USA
20. Department of Neurology, Drexel University College of Medicine, Philadelphia, PA, 19102, USA
21. Department of Neurology, Temple University, 7602 Central Ave, Philadelphia, PA, 19111, USA
22. Department of Neurology, University of Helsinki, Helsinki, FIN-02900, Finland

23. Department of Neurology, Veterans Affairs Boston Healthcare System, Boston, MA, 02130, USA
24. Department of Neurology, University of Massachusetts Medical School, Worcester, MA, 01605, USA
25. Department of Geriatrics, Neurosciences and Orthopedics, Center for Geriatric Medicine, Catholic University of Sacred Heart, Rome, 00168, Italy
26. INM, INSERM, Univ. Montpellier, CHU Nimes, Nimes, France
27. Neuromuscular Division and ALS Center, Beth Israel Medical Center, Albert Einstein College of Medicine, New York, NY, 01605, USA
28. Department of Neurology, Johns Hopkins University, Baltimore, MD, 21287, USA
29. Department of Pathology, Haartman Institute/HUSLAB, University of Helsinki and Folkhalsan Research Center (LM), Helsinki, FIN-02900, Finland
30. Department of Clinical and Movement Neurosciences, UCL Queen Square Institute of Neurology, University College London, London NW3 2PF, UK
31. Department of Medicine (Neurology), Djavad Mowafaghian Center for Brain Health, University of British Columbia, Vancouver, V6T1Z3, Canada
32. Department of Neuroscience, Experimental Neurology and Leuven Research Institute for Neuroscience and Disease, University of California San Diego, 9500 Gilman Drive, La Jolla, CA, 92093, USA
33. Tanz Centre for Research of Neurodegenerative Diseases and Division of Neurology, University of Toronto, Toronto, M5T 0S8, Canada
34. Faculty of Human and Medical Sciences, University of Manchester, Manchester, M13 9PT, UK
35. Neurodegenerative Diseases Research Unit, Laboratory of Neurogenetics, National Institute of Neurological Disorders and Stroke, Bethesda, MD, 20892, USA
36. Department of Neurology, Institute for Clinical Neurobiology, University of Würzburg, Würzburg, D-97078, Germany
37. Department of Neurology, Penn State College of Medicine, Hershey, PA, 17033, USA
38. Genetics and Target Identification, Cerevel Therapeutics, Cambridge, MA 02141
39. Neuromuscular Diseases Research Section, Laboratory of Neurogenetics, National Institute on Aging, Bethesda, MD, 20892, USA
40. Clinical and Neuropathology Core, Johns Hopkins University, Baltimore, MD, 21287, USA
41. KU Leuven – University of Leuven, Department of Neurosciences, Experimental Neurology, and Leuven Brain Institute (LBI), Leuven, Belgium
42. University Hospitals Leuven, Department of Neurology, Leuven, Belgium.
43. Center for Neurodegenerative Disease Research, Department of Pathology and Laboratory Medicine, Perelman School of Medicine at the University of Pennsylvania, Philadelphia, Pennsylvania, USA
44. VIB, Center for Brain & Disease Research, Laboratory of Neurobiology, University of Leuven, Leuven, 3000, Belgium
45. Division of Neurology, Sunnybrook Health Sciences Centre, University of Toronto, Toronto, M4N 3M5, Canada

### Supplementary note 3

#### Consortium acknowledgements

JPR received a doctoral student fellowship from the ALS Society of Canada and a Canadian Institutes of Health Research (CIHR) Frederick Banting & Charles Best Canada Graduate Scholarship (FRN 159279). This work was supported in part by the Intramural Research Program of the National Institute on Aging, National Institutes of Health, Department of Health and Human Services (project ZIA AG000949). Project MinE Belgium was supported by a grant from IWT (n° 140935), the ALS Liga België, the National Lottery of Belgium and the KU Leuven Opening the Future Fund. R.J.C. is funded by the Malta Council for Science & Technology, the Anthony Rizzo Memorial ALS Research Fund and the University of Malta Research Trust (RIDT).
